# Perceptions, attitudes, and challenges regarding continuing professional development among Ethiopian Medical Laboratory professionals: A mixed-method study

**DOI:** 10.1101/2024.02.04.24302318

**Authors:** Adugna Negussie, Abay Sisay, Agumas Shibabaw, Edosa Kebede, Firehiwot Tesfaye, Gizachew Kedida, Nadew Deneke, Takele Teklu, Waqtola Cheneke

**Affiliations:** Department of Medical Laboratory Sciences, College of Health Sciences, Arsi University, Asella, Ethiopia; Department of Medical Laboratory Sciences, College of Health Sciences, Ababa University, Addis Ababa, Ethiopia; Department of Medical Laboratory Sciences, College of Medicine and Health Sciences, Wollo University, Dessie, Ethiopia; Department of Medical Laboratory Sciences, College of Medicine, and health Sciences, Ambo University, Ambo, Ethiopia; Department of Medical Laboratory Sciences, College of Medicine, and health Sciences, Dire Dawa University, Dire Dawa, Ethiopia; Ethiopian Medical Laboratory Association, Addis Ababa, Ethiopia; School of Medical Laboratory Science, College of Health Sciences and Medicine, Wolaita Sodo University, Wolaita Sodo, Ethiopia; School of Medical Laboratory Science, Faculty of Health Science, Institute of Health, Jimma University, Jimma, Ethiopia

**Keywords:** attitude, CPD, medical laboratory professionals, perception

## Abstract

**Background:** Medical laboratory professionals play vital role in healthcare. The growing demand for quality laboratory services and emerging technologies underscore the crucial need for Continuing Professional Development (CPD). However, there is limited information on CPD programs in Ethiopia. Thus, this study aimed to assess medical laboratory professionals’ perceptions, attitudes, and challenges towards CPD and improve engagement to enhance diagnostic service quality.

**Methods:** this cross-sectional study enrolled 228 medical laboratory professionals in Ethiopia from July to October 2023. Using a mixed-methods approach that combined quantitative data from an online survey and qualitative data from interviews. SPSS version 28 was used for data analysis.

**Results:** the average age of the study participants was 32.6 ±6.4 (SD) years, the majority were men (88.6%), and 44.3% have worked for more than ten years. Of the participants, 51% never had CPD training. About three-fourth of the participants perceived CPD as essential to their professional career. About 45.2% of the study participants perceived that the purpose of CPD course is to renew their license and gain knowledge and skills that are not covered in basic training. While the majority of participants had good attitudes towards CPD, about 10% of them stated that it is not important in their career growth. The majority of the study participants were not in support of the decentralized CPD system. A notable problem with finance, insufficient manpower, unsupportive employers, a lack of awareness by regulatory bodies, inadequate access to training close to their working area, were identified as significant challenges of the CPD program.

**Conclusion:** the study highlights the perception that CPD is crucial for enhanced laboratory practices and career advancement. The study highlights the need for targeted strategies to address the identified problems and increase the engagement of medical laboratory professionals in the CPD program.

## Introduction

Medical laboratory professionals are responsible for modern healthcare. They conduct complex scientific procedures on a range of biological materials using conventional and cutting-edge technologies, contribute to accurate diagnoses, effective treatments, disease prevention and control. They play vital role in the advancement of medical practices in influencing the policy [1, 2]. The growing demand for quality laboratory services and emerging technologies underscore the crucial need for medical laboratory professionals to continuously enhance their expertise. This can be ensured through effective continuous professional development (CPD) programs [3, 4].

CPD involves various learning activities to maintain and enhance skills of health professionals. It enables safe, effective, and legal practice in a changing scope of practice [5]. CPD is crucial for healthcare workers, helping them maintain and enhance their competence throughout their careers. It ensures they stay updated with current practices and advancements in the industry [6]. CPD also supports career development, allowing professionals to acquire new skills and knowledge [7]. Medical laboratory professionals need to stay informed and enhance their expertise [7, 8].

Moreover, CPD has become a mandatory requirement for health professionals in Ethiopia, like in many other countries, for re-licensure purposes to practice in the healthcare system, and provide patients with the highest level of care [8, 9]. This may ultimately lead to better care and health outcomes [10].

In low and middle-income countries, including Ethiopia, taking part in CPD has been challenging for healthcare professionals due to various obstacles. These include limited access, lack of time, insufficient compensation, and inadequate awareness of CPD [8, 11, 12, 13]. Financial cost and workload have also been identified as key barriers to participation in CPD courses [9,14]. Additionally, there are problems with course coverage, the inability to take paid or unpaid study leaves, understaffing, and leadership issues that hinder healthcare professionals from attending CPD programs [15].

Ethiopia is the second most populous country in Africa, with a population of more than 110 million. The country characterized by low socioeconomic status and health services. It has about 25,517 public and private health facilities of which more than 7,000 have laboratories as diagnostic health facilities [16]. However, there is a low engagement of medical laboratory professionals in the existing CPD program [17]. Published information in this regard is very limited in Ethiopia; thus, this study aimed to assess the perception, attitude, and challenges of medical laboratory professionals towards CPD. By identifying the barriers, policymakers, professional bodies, and other concerned stakeholders can design appropriate strategies to promote and enhance medical laboratory professionals’ engagement in CPD activities and ultimately improve the quality of diagnostic service provided to patients.

## Materials and Methods

### Study Setting, Period, and Participants

This study was carried out in Ethiopia from July to October 2023, aiming to assess the perception, attitude and challenges on CPD. A total of 228 medical laboratory professionals who were working in 12 regions and two city administrations of the country were recruited for this study.

Medical laboratory professionals working in public, private, and non-governmental organizations (NGOs) that offer medical laboratory services, were approached through academic and university-referral hospitals, clinics, and regional reference laboratories to participate in the online survey. In addition, for the qualitative part, 20 study participants with various academic backgrounds and currently working in CPD related positions were recruited as key informants for an in-depth interview. The information obtained from study participants can give a clue about the perception and attitude of medical laboratory professionals towards CPD and its role in improving laboratory services and designing a solution for CPD-related challenges.

### Study Design and Sample Size

A cross-sectional online survey was conducted using a structured self-administered Google-based data collection tool. The study used a mixed-method approach, combining both quantitative and qualitative data collection methods. The sample size was based on the willingness of the respondents to the online survey. Accordingly, 228 professionals responded to the online survey. To triangulate the quantitative data, we used qualitative data from an in-depth interview of purposely selected CPD centers and regulatory bodies. The study intends to explore how Ethiopian medical laboratory professionals perceive opportunities for continuing professional development, their attitudes, and challenges that may hinder their attendance.

### Data Collection and Quality Assurance

A structured self-administered online data collection tool was used to collect the quantitative and qualitative data. A Google survey link was shared, allowing them to conveniently respond. Study coordinators were assigned in the respective regions to facilitate the data collection. The online data collection tool developed for this survey had four sections: demographics, professional perception, and attitude towards CPD, challenges of attending CPD, and cross-cutting issues. The tool was developed specifically for medical laboratory professionals and was pre-tested for accuracy and consistency prior to the actual data collection. Each area of the perception tool was scored using the three-point Likert scale as 1 = strongly agree; 2 =agree and 3 = disagree. The study also employed a qualitative data collection tool consisting of semi-structured interviews with 20 respondents from selected institutes. These in-depth interviews were used to gather additional insights and information that could not be captured through the online survey. The data collection tool was pre-tested for its accuracy and consistency. Completion, accuracy, and clarity of the collected data were checked carefully and regularly.

### Data Analysis and Interpretation

The quantitative data were analyzed using SPSS version 28, with descriptive analysis (frequency, percentage, mean, range), and 3-point Likert data was used to describe the perceptions and attitude of the laboratory professionals on CPD. The top priority of CPD-related challenges and specific issues were identified using mean distributions. The qualitative data were triangulated with the online collected quantitative data for enriching and linking the identified gaps and analyzed by themes.

Attitude was measured by mean score level. Above the mean scores were considered as good attitude.

### Ethical considerations

Ethics approval was obtained from Ambo University, College of Medicine, and Health Sciences before the commencement of this study. Participants were asked for their consent prior to enrollment. All information was acquired and reviewed in an anonymous method to protect participant privacy; no form of personal identification was included in the data collection instrument. The data will only be used by the association to guide the intervention and evaluate its success.

## Result

### Characteristics of Study Participants

A total of 228 medical laboratory professionals participated in this online survey. The mean age of the participants was 32.6±6.4 (SD) years and ranged from 22 to 55 years. The participants had varying years of service, ranging from less than one year to over 10 years of experience. The majority of the participants were males 88.6% (202/228), from government hospitals (41.6%), had bachelor’s degrees in medical laboratory science/Technology (MLS/T) (55.7%), and more than ten years of experience (44.3%). Fifty-one (116/228) percent of the participants never take CPD training. The sociodemographic characteristics of the study participants are summarized in **Table 1**.

**Table 1:** Characteristics of study participants (N = 228)

| **Characteristics** | **Frequency** | **Percentage** |
| --- | --- | --- |
| **Sex** |  |  |
| Female | 26 | 11.4 |
| Male | 202 | 88.6 |
| **Age (in year)** |  |  |
| 21–30 | 103 | 45.2 |
| 31–40 | 103 | 45.2 |
| >40 | 22 | 9.6 |
| **Participants Region** |  |  |
| Oromia | 75 | 32.9 |
| Addis Ababa | 40 | 17.5 |
| SNNPR* | 38 | 16.7 |
| Dire Dawa | 13 | 5.7 |
| Sidama | 12 | 5.3 |
| Tigray | 12 | 5.3 |
| Gambelia | 10 | 4.4 |
| Afar | 7 | 3.1 |
| Benishangul Gumz | 7 | 3.1 |
| Harari | 6 | 2.6 |
| Somali | 4 | 1.8 |
| Amhara | 2 | 0.9 |
| South-West Ethiopia | 2 | 0.9 |
| **Educational Level** |  |  |
| BSc | 127 | 55.7 |
| MSc | 80 | 35.1 |
| Diploma/level IV | 18 | 7.9 |
| PhD | 3 | 1.3 |
| **Years of service** |  |  |
| <1 | 19 | 8.3 |
| 1–5 | 46 | 20.2 |
| 6–10 | 62 | 27.2 |
| >10 | 101 | 44.3 |
| **Participants' current place of work** |  |  |
| Government Hospital | 95 | 41.6 |
| Academic institute, University/ College | 37 | 16.2 |
| Regional Laboratory | 36 | 15.8 |
| Government Health Center | 26 | 11.4 |
| NGO | 12 | 5.2 |
| Research Institute | 6 | 2.6 |
| Private academic institutions and health facilities | 11 | 4.8 |
| Other | 5 | 2.2 |
| **Ever attend CPD training** |  |  |
| No | 116 | 50.9 |
| Yes | 112 | 49.1 |

\*SNNPR= South Nations Nationalities People Region (current South and Central Ethiopia)

### Perception of study participants on the importance of CPD

In the current study, 76.8% (175/228) of the participants believed that CPD is important for their professional careers (**Table 2**). This finding was also supported by the qualitative study; one participant responded to the importance of CPD as; “it is good but does not accommodate/reach all professionals who work in remote areas and the training should cover all practical areas and updated contents of CPD program”. Another respondent also described that “CPD is a good idea, but training opportunities are very low, it would be good if online courses were also available”. Among the different CPD directors and regulators, a participant stated that “CPD is a critical component of human resource development to provide quality health care by competent health professionals”, another participant added that “CPD is very important if professionals understand it as this is for their benefit to update their knowledge and skill”. In addition, one of the participants explained the importance of CPD by his statement “I see CPD as a building block for health care workers’ competency and practicing safely across the nation”. However, 17.1% of the participants did not have any say about CPD and 6.1% of participants perceived that CPD is less important.

**Table 2.** Perception of study participants on the importance of CPD.

| Response of participants | Frequency | Percent |
| --- | --- | --- |
| CPD is important | 114 | 50.0 |
| CPD is very important | 61 | 26.8 |
| No opinion | 39 | 17.1 |
| Not that match important | 14 | 6.1 |
| Total | 228 | 100.0 |

About 45.2% of the study participants perceived the need for a CPD course is to gain knowledge and skills not obtained during the basic training and to renew their license. About 43.4% perceived CPD as important to renew their license. While the remaining needed the course as stated in **Table 3**.

**Table 3.**
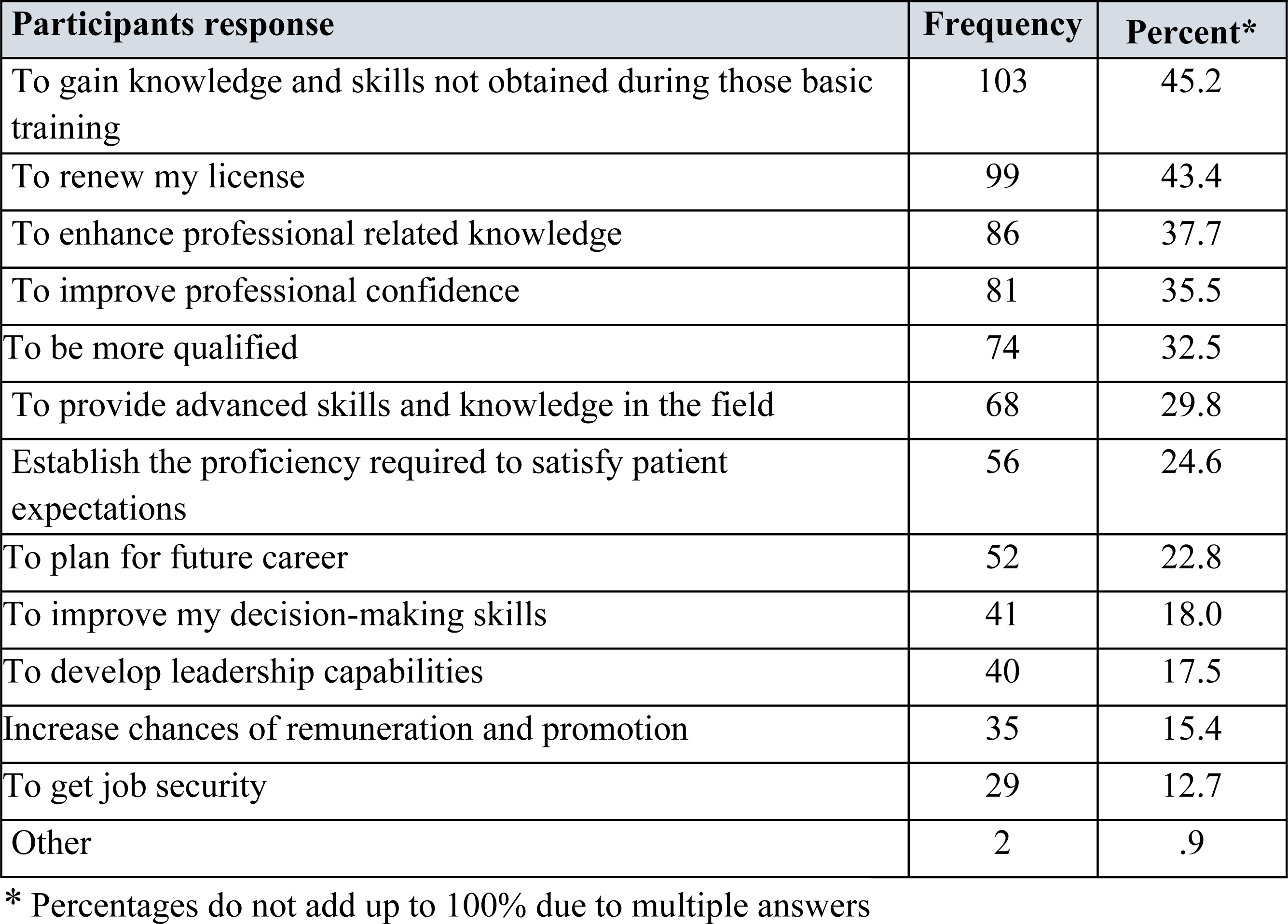
Perceptions of study participants on the purpose of CPD.

### Perception of study participants toward decentralized CPD program

Nearly 60 percent of the study participants did not support the decentralized provision of CPD courses for professionals (**Figure 1).** Most professionals believe that this way of the CPD program compromises the quality and goal of the program, and it is also enhancing illegal certification.

**Fig 1.**
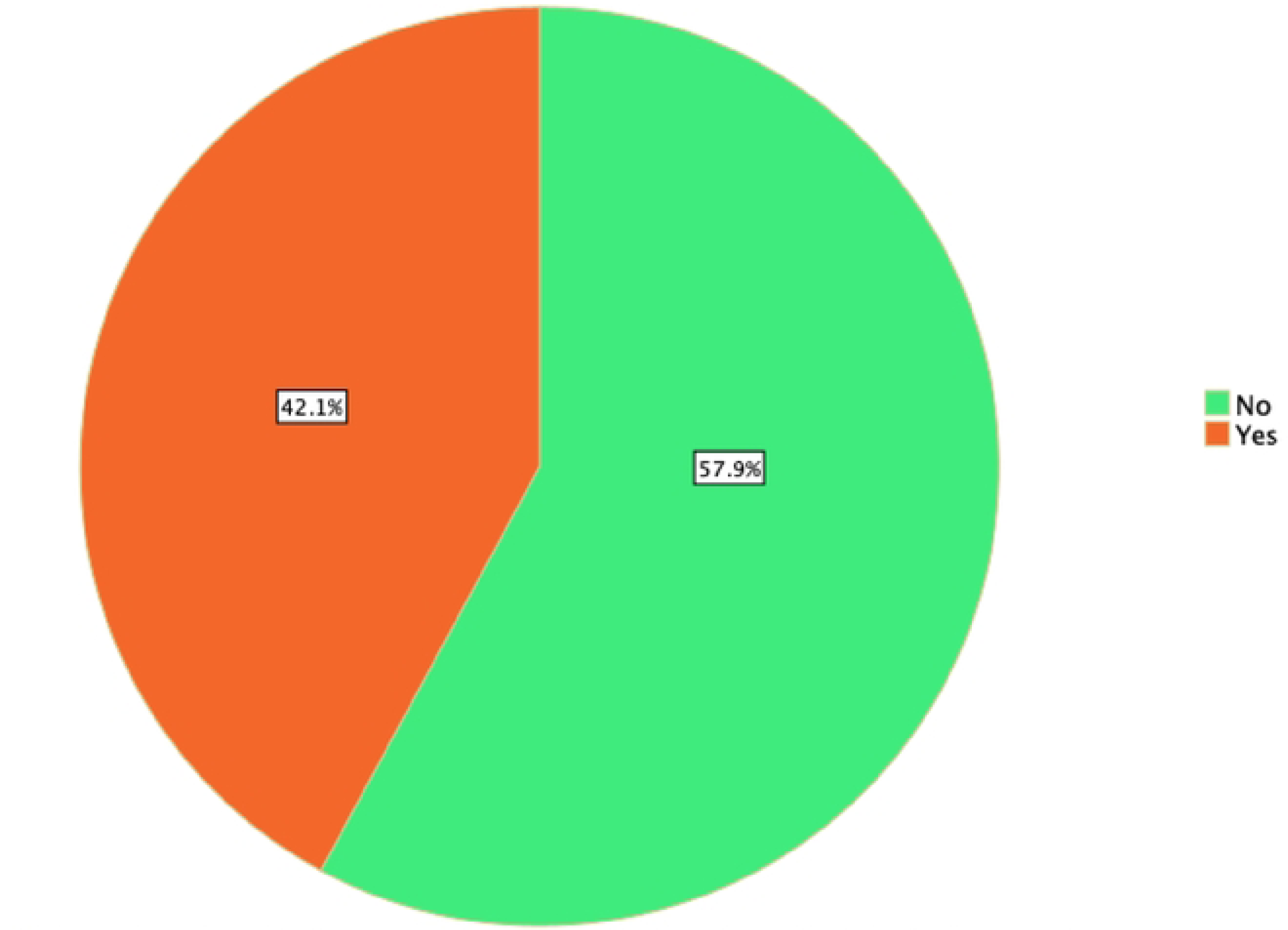
Perception of study participants on highly decentralized CPD provision.

Respondents in their open questionnaires describe the situation of decentralized CPD as follows:

“…the quality of training materials, trainers’ education level, training facilities will be compromised and may shift the objectives of the CPD training to commercial aspect”,

“…the training quality might be affected and compromised by substandard training sessions, in terms of training modality, trainer competency, and the delivery methods, …”,

“It became subjective”, and “because it mainly focuses on CPD for license renewal rather than developing basic skills the profession requires”,

“Current policy affects the intention of CPD because everybody needs it for license issues and this will affect or enforce them to compromise the quality, this CPD must be given for free and authorized by government otherwise private sector may affect the quality since their focus is mainly to make a profit”,

### Attitude of study participants towards CPD

Most of the participants in this study strongly agreed that CPD is a learning activity after basic formal education (75.9%), is a process of lifelong learning for individuals and teams (74.1%), involves keeping up to date with knowledge and skills (81.6%), is a process that supports flexible career pathways and help career aspiration (73.7%), involves participation in workshops, seminars, conferences, journaling, research (69.3%) as well as is important and mandatory for the medical laboratory professionals (79.4%). Conversely, few of the participants disagreed that CPD involves keeping up to date with knowledge and skills (5.7%) while relatively fewer participants also disagreed that CPD is a process that supports flexible career pathways and help career aspirations (7.9%) as well as involves participation in workshops, seminars, conferences, journaling, and research (8.8%) as shown in **Table 3** and **Figure 2**. Approximately 10% of participants explained that CPD is not important and mandatory for laboratory professionals (Table 3-6).

**Fig 2.**
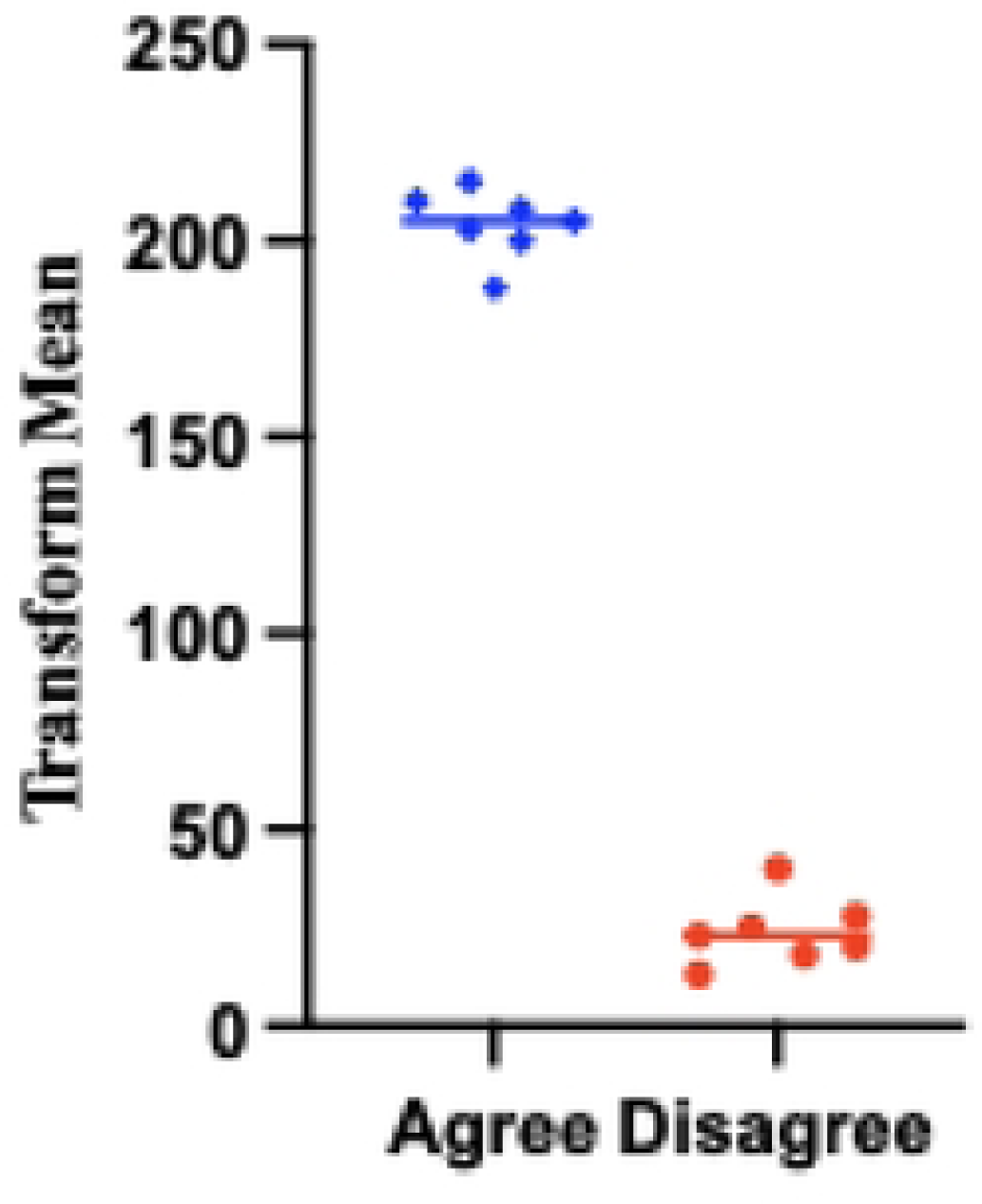
Transformed data of study participants’ attitude towards CPD.

### Satisfaction of study participants on the existing training

We also tried to assess how satisfied the participants were with the existing CPD training. Accordingly, only 55.4% of them were satisfied with the trainer qualification and experience. The survey showed that under half of the study participants attending the training were satisfied with the quality of the training module, and about a quarter of them on training modality, training facility, and refreshments as stated in Table 5.

**Table 4.** Attitude of study participants towards CPD.

| **Participants response** | **Agree**<br>**N (%)** | **Disagree**<br>**N (%)** |
| --- | --- | --- |
| CPD involves engaging in formal training after basic training only | 188 (82.5) | 40 (17.5) |
| CPD is a learning activity after basic formal education | 203 (89.0) | 25 (11.0) |
| CPD is a process of lifelong learning for individuals and teams | 200 (87.7) | 28 (12.3) |
| CPD involves keeping up to date with knowledge and skills | 215 (94.3) | 13 (5.7) |
| CPD is a process that supports flexible career pathways.<br>and help career aspirations | 210 (92.1) | 18 (7.9) |
| CPD involves participation in workshops, seminars,<br>conferences, journaling, research | 208 (91.2) | 20 (8.8) |
| CPD is important and mandatory for the Medical Laboratory<br>Professionals | 205 (89.9) | 23 (10.1) |

**Table 5.** Satisfaction of study participants on the existing CPD training.

| Participant response | Frequency | Percent |
| --- | --- | --- |
| Trainer qualification and experience | 62 | 55.4 |
| Module quality | 42 | 37.5 |
| Training quality | 28 | 25.0 |
| Training modality | 26 | 23.2 |
| Refreshment | 25 | 22.3 |
| Training facility | 24 | 21.4 |
| Other | 6 | 5.4 |

### CPD training modalities suggested by study participants

More than three-fourths of the participants suggested a face-to-face approach for CPD courses, while the online and workshop approaches were also suggested as presented in **Figure 3**.

**Fig 3.**
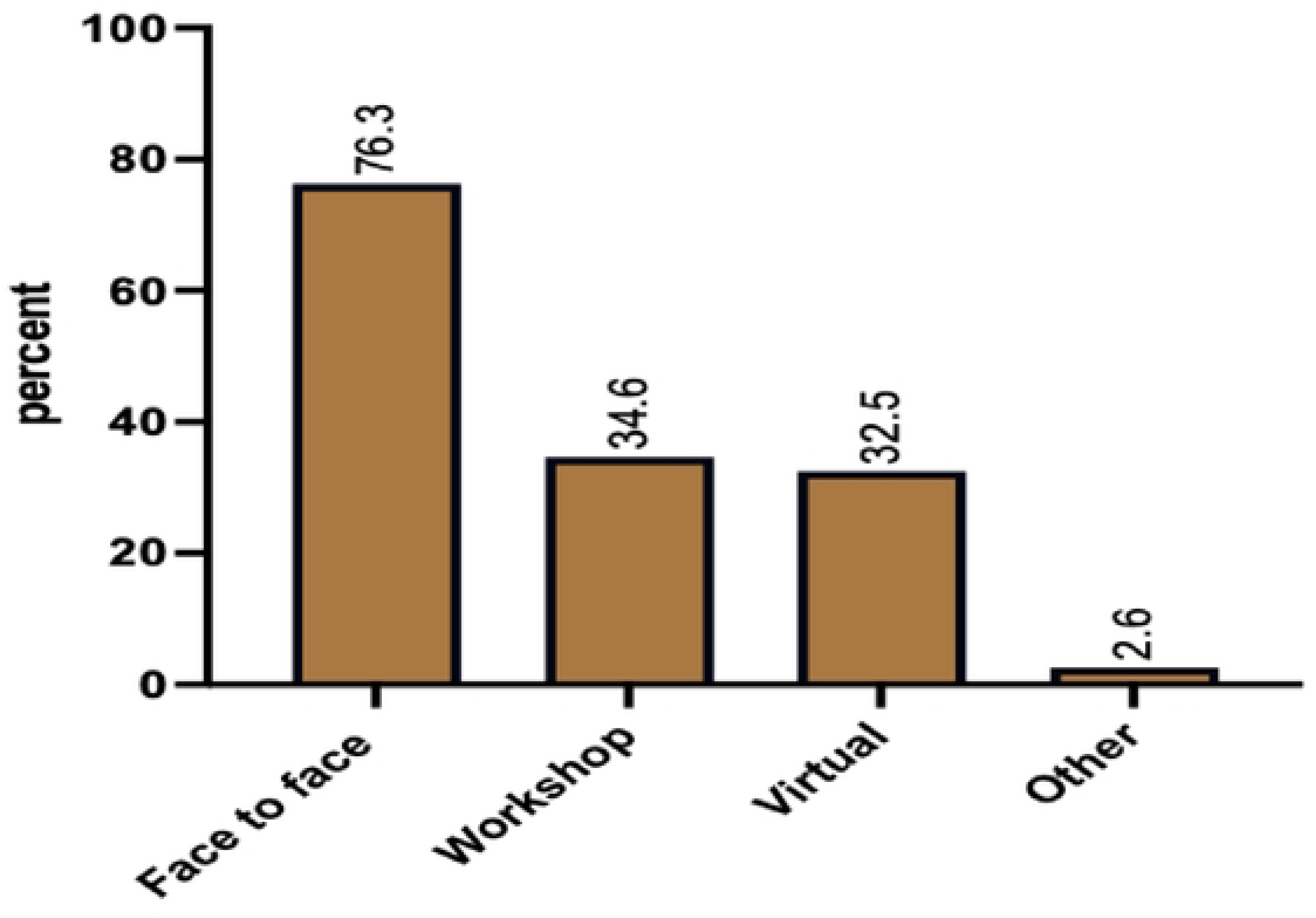
Types of 1nodalities for CPD training suggested by study partici1>ants.

### Challenges in CPD Program

The identified challenges to the CPD program were increased cost for CPD training, low access for training, limited information or awareness, and minimal employer support. Among these, lack of funds or financial support was reported as the most significant challenge by the majority of study participants, with 68% of them reflecting on this issue. Related to CPD course cost participants described their perception as; “the cost for CPD does not consider the income of professionals”, “it does not take into context the financial capability of health professionals’’, “I think it’s very exaggerated”, “it should take into consideration the income of government employees”, “EMLA’s CPD cost is expensive compared to others”, “it should be free for EMLA members’’, “online CPD is better than face to face as no need of other budgets”, and “governmental and even private health institutions should consider covering the expenses for courses”.

The limited availability of accredited CPD courses in a face-to-face and online modality was another challenge that was reflected by study participants. Respondents described that there is a lack of relevant learning opportunities due to a scarcity of learning materials in their proximity. One of the study participants described that there is “no laboratory-related training module and center to take the training”.

Professionals have raised concerns about the distance of training centers, considering it as a significant challenge. They believe that their remote location acts as a barrier and there aren’t enough local learning opportunities. In addition, some private organizations have monopolized CPD, and the training of trainers (TOT) was not cascaded to lower-level facilities. Another respondent pointed out that CPD training does not consider the demographic information of the health facility and the training need assessment.

Respondents also point out the lack of employer support as an additional barrier to implementing CPD of these: “lack of commitment in upper management”, “heads of the facility or department are not willing to send their staff to CPD training”, “private institutes are not cooperative” and “lack of government and regulatory support”. In addition to this, there is a gap among the professionals in supporting each other or networking and overall awareness of CPD. This is supported by the respondent’s description of; “lack of commitment from the professionals”, “lack of awareness” and “poor networking”. Table 6 details the challenges of the study participants towards the CPD program.

**Table 6.** Identified challenges by study participants on CPD implementation.

| Participant response | Frequency | Percent |
| --- | --- | --- |
| Lack of funds for attending the training | 155 | 68.0 |
| Lack of relevant learning opportunities | 77 | 33.8 |
| Lack of employer's co-operation to allow time away from the workplace | 65 | 28.5 |
| Shortage in time | 65 | 28.5 |
| Lack of sufficient information on the training center | 51 | 22.4 |
| Lack of administrative cooperation | 35 | 15.4 |
| Lack of early notification of the program | 33 | 14.5 |
| CPD training center is far from my working place | 29 | 12.7 |
| Scheduled for the program is inappropriate | 26 | 11.4 |
| Lack of interest in topic or module | 26 | 11.4 |
| Lack of quality training | 24 | 10.5 |
| Shortage of staff | 10 | 4.4 |
| Negative experiences with previous CPD program | 7 | 3.1 |
| Other | 1 | 0.4 |

According to qualitative interviews with key informants from CPD centers and regulatory organizations, the study found that there is no formal regulatory framework in place to oversee the entirety of the courses offered as part of the CPD program. Taking the required credit hours is currently emphasized by the regulatory authority, regardless of the certificate’s source or significance in specific professions. They point out that there are currently no standards for the caliber of the CPD program. According to one key informant, participants “targeted on license renewal without considering whether professionals had really gained new knowledge and skill or not”. Another participant from the regulatory authority also explained how they prioritize CEU, the year the course is offered, the kind and degree of profession studied, and regardless of the relevancy of the course.

## Discussion

This was the first cross-sectional mixed-method study conducted in Ethiopia to assess the perception, attitude, and challenges toward CPD implementation among medical laboratory professionals in an effort to implement CPD as an organized method of learning and professional development. Assessing the perception, attitude, and challenges of professionals is crucial in developing a comprehensive CPD program that enhances their competencies. The results of the survey showed that, in addition to helping them renew their licenses, they understood that CPD is crucial for their professional career. Although respondents linked CPD to professional competence, there is limited evidence to support the supposition that undertaking regular CPD leads to the advancement of professional competence.

The majority of professionals in the current study were aware of the value of continuing professional development (CPD) in advancing their careers. Nonetheless, less than 50% of the laboratory personnel surveyed reported having attended any of the current CPD courses. The results demonstrated that, in comparison to other professionals in the nation, there is a low level of overall involvement in CPD programmed. This discrepancy in CPD attendance may be caused by professionals’ lack of understanding of its implementation, limited access to courses, uncertainty about the caliber of the dispersed training provided elsewhere, and/or upper management’s commitment to the adoption in the healthcare system. Above that, there are no specific guidelines that dictate how health professionals with specific fields can practice in the implementation of CPD.

This study indicated that there is a gap in attitude between laboratory professionals towards CPD. This indicates that best practices in laboratory professions education have not been universally translated into CPD practice, and the development and implementation of these strategies vary among different contexts and institutions. That is why unexpected findings were recorded from professionals and should require more advocacy, perception, and behavioral change towards CPD.

The majority of research participants expressed interest in in-person or face-to-face training options and being near their places of employment. They demonstrated the necessity of the theoretical and practical course materials, which are promoted in CPD courses. Furthermore, they indicated the necessity for several free or inexpensive online continuing education courses. The finding of this study also showed the need for a centrally authorized CPD course in the country to avoid confusion on the goal of the CPD program. The outcomes align with a University of Botswana study about the same occupation [18].

The survey assesses various challenges for the low achievements in the implementation of CPD for laboratory professionals. The main challenges encountered by respondents was a lack of financial resources, limited course access, distance of the training center, lack of time, and low leaders’ support are among the most common factors challenging professionals for their engagement in CPD program. This is consistent with research on continuing professional development (CPD) for other professionals elsewhere; radiologists [19] nurses [20] and pharmacists [21]. In addition, there is a scarcity of workers to cover during the training when it is conducted outside the center, inadequate information about the CPD course, and late notification. Qualitative data from the current study showed that professionals working in remote places had faced challenges related to travel distance and time for scheduled CPD training, as well as limited peer support networks and face-to-face meeting possibilities.

There is no reasonable resource allocation for CPD program by the Ministry of health, regional health bureaus, and health facilities (both public and private) to create a suitable platform of CPD access for professionals except for some disease programs with external funding. There is also no strong regulation pertaining to the responsibilities of employers to facilitate CPD training for their employees. In addition to this regulatory bodies are not checking CPD certificates presented by professionals during license renewal whether the certificates are genuine, and whether the courses are relevant or not despite there being overwhelming evidence of fraud and irrelevant courses taken by healthcare professionals in general and laboratory professionals in particular. Regulatory bodies further claim that they are not mandated to check for certificate authenticity or relevance of CPD courses.

It is also known that there is a clear lack of information and awareness on the requirements of national guidelines for the CPD implementation and participation by regional, facilities management personnel, regulatory bodies and professionals contributing to the hindrances of the implementation. Many of the laboratory managers are not aware of the CPD courses accreditation requirements, and they don’t inquire their hospital managers for needed appropriate CPD courses for their subordinates, while institutional management also don’t worry about the needed resources’ allocation for the training of laboratory professionals.

The dynamic growth nature of medical laboratory science has made it necessary for laboratory professionals to update their skills and abilities on a regular basis. It is recommended that a variety of continuing professional development programs be made available on a regular basis, as this will be an important step in efforts to improve the provision of quality health care. The cost of the CPD courses should be revised or to take stakeholder support into account, and a centrally accredited program that will be offered at a location of professional’s choice or close to their workplace will be necessary rather than the highly decentralized CPD implementation system which may compromise quality of services. To achieve the desired outcome and ascertain how the program influences staff competency, efficient monitoring and evaluation of the program should be put in place. Additionally, we advise the MOH to work with the professional association and other concerned bodies to implement such programs to raise the level of expertise among staff members in medical facilities, which will enhance patient care and the standard of medical service. Professional associations should be authorized to play a central role in the CPD implementation specifically in the accreditation of CPD facilities and courses for their respective profession, and they should also be given the responsibilities to authenticate CPD certificates before license renewal by regulatory bodies. Furthermore, we recommend that professional associations should develop specific CPD implementation guidelines for their respective professions and be mandated to oversee the ongoing processes of the CPD with their respective professional members.

### Strength of the study

This was the first study on the perception of CPD that considered Ethiopian medical laboratory professionals. The study tried to triangulate quantitative data by using the qualitative from key informants selected from the regional and Ministry of Health CPD directors and coordinators. This research is regarded as an opportunity to introduce the concept of CPD and demonstrate how laboratory professionals perceive this new approach that maintains and keeps professionals updated on evidence-based practices and recommendations, which are also a crucial part of maintaining patient safety culture and safe patient care. Furthermore, this study may encourage educators, researchers, policymakers, and laboratory professionals to reconsider the extent to which a program of this kind is necessary, mandated, and appropriate to various health institutions.

### Limitations of the study

There are certain limitations on this study. For instance, most of the information was collected via an online survey and did not include observational study. Due to the security problem and restricted internet access, it was difficult to reach out to participants from the Amhara region.

## Conclusion

The majority of participants felt that CPD was crucial to their professional careers, despite the fact that professional involvement in CPD has decreased. The majority of research participants did not favor the decentralized CPD strategy. They thought that CPD ought to be mandated as it offers a great chance to improve laboratory practices, map out career paths, stay up to date on professional advancements, increase self-confidence, enhance opportunities for promotion, and provide with knowledge and skills that did not receive in basic training. A professional engagement in ongoing CPD may be hampered by components such as lack of funding, time restraints, limited access for training close to their working place, an unsupportive work environment, and their employment situation.

## Data Availability

All relevant data are within the manuscript and its Supporting Information files.

## Acknowledgments

First, we would like to thank the participating Ethiopian medical laboratory professionals for their invaluable time and information shared for this survey. Second, we express our gratitude to Professor Aster Tsegaye for her critically reviewing this work. Lastly, we would like to express our gratitude to EMLA for providing the necessary support for this study over its whole duration, and USAID/Jhpiego for financial support of the project.

## Author Contributions

Conceptualization and investigation: Adugna N, Abay S, Agumas S, Edosa K, Firehiwot T, Gizachew K, Nadew D, Takele T, Waqtola C.

Methodology drafted: Abay S, Firehiwot T.

Software data collection and formal analysis: Adugna N.

Writing – original draft Background and justifications: Edosa K, Nadew D, Waqtola C

Writing – original draft result and discussion: Adugna N, Agumas S

Writing – review & editing: Adugna N, Abay S, Edosa K, FirehiwotT, Gizachew K, Nadew D, Takele T, Waqtola C

Validation: Adugna N, Abay S, Agumas S, Edosa K, Firehiwot T, Gizachew K, Nadew D, Takele T, Waqtola C

Project administration: Adugna N and Gizachew K

## Data Availability Statement

All data sets used in this study are included in this manuscript.

